# Improvement of field-deployable metagenomic virus detection by a simple pretreatment method

**DOI:** 10.1101/2021.12.31.21268552

**Authors:** Anna S. Fomsgaard, Morten Rasmussen, Katja Spiess, Anders Fomsgaard, Graham J. Belsham, Jannik Fonager

**Author notes:** **Corresponding Author:** Anna S. Fomsgaard **Corresponding E-mail:**. **Mail addresses:** Anna Fomsgaard, Morten Rasmussen, Katja Spiess, Graham Belsham, Anders Fomsgaard, Jannik Fonager.

## Abstract

As both the current COVID-19 pandemic and earlier pandemics have shown, animals are the source for some of the deadliest viral pathogens, which can spread to humans. Therefore, early detection at the point of incidence is crucial to both prevent and understand the threats posed to human health from pathogens in animal reservoirs. Often, the exact genetic nature of these zoonotic pathogens is unknown and advanced laboratory facilities do not exist in most field settings and therefore the development of methods for unbiased metagenomic and point of incidence detection is crucial in order to identify novel viral pathogens in animals with zoonotic and pandemic potential.

Here we addressed some of these issues by developing a metagenomic Nanopore next-generation sequencing (mNGS) method for nucleic acids extracted from clinical samples from patients with SARS-CoV-2. To reduce the non-RNA viral genetic components in the samples, we used DNase pretreatment in a syringe followed by filtration and found that these pretreatments increased the number of SARS-CoV2 reads by > 500-fold compared with no pretreatment.

The simple protocol, described here, allows for fast (within 6 hours) metagenomic detection of RNA viruses in biological samples exemplified by SARS-CoV-2 detection in clinical throat swabs. This method could also be applied in field settings for point of incidence detection of virus pathogens, thus eliminating the need for transport of infectious samples, cold storage and a specialized laboratory.

**Highlights:**

- Here we present a field-deployable metagenomic Nanopore protocol for RNA virus detection.
- SARS-CoV-2 detection used as proof-of-concept.
- Analysis of simple methods for non-viral nucleic acid depletion were tested on SARS-CoV-2 clinical samples.
- DNase I treatment followed by 0.22µM syringe filtration dramatically increased the number of SARS-CoV-2 reads and virus genome coverage.

## 1. Introduction

SARS-CoV-2 was first identified in China in late 2019 (Wu et al., 2020) and by early spring 2020 the infection had developed into a pandemic; the virus has infected over 250 million people and caused over 5 million deaths at the time of writing (WHO, 2021). The SARS-CoV-2 has a positive-sense single-stranded RNA genome of app. 30,000 nucleotides (nt) that is prone to accumulate mutations, which may occasionally facilitate a host species jump. Though its precise origin is unknown, closely related coronaviruses have been found in bats (H. Zhou et al., 2020a; P. Zhou et al., 2020b). In addition to the human infections, SARS-CoV-2 infections have been detected in several animal species including American mink, bats, felines (like domestic cats, tigers, lions), white-tailed deer and canines (Garigliany et al., 2020; Hammer et al., 2021; Kuchipudi et al., 2021; Larsen et al., 2021; McAloose et al., 2020; Munnink et al., 2021). These diverse host infections highlight the ability of zoonotic RNA viruses like SARS-CoV-2 to cause outbreaks, even pandemics. Areas with high wildlife biodiversity, deforestation and urbanization are especially vulnerable to such serious events (Gardy & Loman, 2018). Therefore, rapid and sensitive identification methods are important to discover and manage new and ongoing threats from viral pathogens including, in some cases, field-detection at the point of incidence.

Metagenomic next-generation sequencing (mNGS) has the ability to identify pathogens in a hypothesis-free manner compared to specific targeted strategies such as antigen or antibody detection, reverse transcription quantitative PCR (RT-qPCR) or other specific nucleic acid (NA) amplification methods (Storch, 2000). Designed primers and/or probes can become obsolete if the pathogen of interest mutates in the primer/probe targeted region, e.g. as observed for SARS-CoV-2 in the S-gene (Ziegler et al., 2020). Additionally, different pathogens can produce similar symptoms, which can lead to multiple independent tests being required to provide a diagnosis. mNGS avoids these limitations since all NA present within a sample are sequenced, resulting in both host and pathogen being detected at the same time (Simner et al., 2018). However, mNGS is not without challenges, e.g. the proportion of the viral genomes can be very small compared to the host-genome, bacterial genomes, plant genomes and other nucleic acids in a given clinical sample (Simner et al., 2018). This “viral needle in a metagenomic haystack” has proven troublesome. In attempts to overcome this problem, two main strategies have been preferred: either virus enrichment or host depletion. Virus enrichment may involve some degree of positive virus selection, e.g. probe capture, poly-A-selection. However, these methods are specifically targeted since probes are virus specific and not all viral RNAs have poly-A tails. In contrast, host depletion involves removing as much non-viral NA as possible. Host depletion can include pretreatment of samples by low-speed centrifugation, ultracentrifugation, filtration, polyethylene glycol precipitation or, depending on the pathogens of interest, the level of DNA or RNA genomes can also be reduced, e.g. using nucleases (Conceição-Neto et al., 2015; Greninger et al., 2015; Hall et al., 2014; Matranga et al., 2014; Sauvage & Eloit, 2016). Although overcoming the metagenomic haystack problem is possible, the workflows can become complicated with many “hands-on” steps. This provides a need for trained personal and specialized laboratories both for wet-lab and bioinformatics analyses and thus, becomes time-consuming and expensive. Some of these complex workflows can be automated, e.g. by utilizing robots for NA extraction. However, considering the ability to make mNGS available for a larger research community and possibly field-deployable, then minimal equipment and fast workflow should be also investigated and prioritized. This may be of major interest in those geographic areas that are potential hotspots for zoonotic pathogen-spillover events.

Here, we present a protocol that includes sample pretreatment, NA extraction, and random-amplification followed by untargeted sequencing using the Oxford Nanopore Technology system for real-time diagnostics. The main objective was to investigate which pretreatments would provide the largest increase in detection of viral NA from clinical swab samples.

## 2. Method

### 2.1. Clinical samples

#### 2.1.1. Pooled samples

Sixteen fresh non-frozen SARS-CoV-2 PCR positive (16 samples with a median CT-value of 22 IQR=7.7) oropharyngeal swab samples were collected at the Test Center Denmark (TCDK), Statens Serum Institut (SSI), Copenhagen, Denmark (SSI, 2021).

#### 2.1.2. Individual samples

Fourteen positive samples (with CT-values in the range 18-36) and a negative sample (with CT>38) were collected between August and September 2021 for pretreatment analysis. All positive samples had been typed to be of the Delta variant B.1.617 using the variant-PCR surveillance program from TCDK screening for the L452R substitution but with the absence of the deletion H69-70 or the N501Y and the E484K substitutions used to type other variants.

#### 2.1.3. Ethics

Exemption for review by the ethical committee system and informed consent was given by the Committee on Biomedical Research Ethics - Capital region in accordance with Danish law on assay development projects. Samples were anonymized prior to analysis.

### 2.2. Pretreatment and extraction

#### 2.2.1 DNase treatment

For DNase treatment, we used the Zymo Research DNase I Set (Zymo Research) according to the manufacturer’s recommendations with 15 minutes incubation at room temperature, except that the volumes were scaled up to 1 ml with PBS.

#### 2.2.2. Filtration

For filtration treatment, we used a 5 ml syringe attached to a 0.22 µM Minisart® Syringe Filter (Sartorius Stedim Biotech, France) to process 1 ml of sample material. An additional 1 ml of air in the syringe ensured that the volume otherwise lost in the filter’s dead-volume was recovered.

#### 2.2.3. Nucleic Acid recovery

NA extraction was performed using the MagNA Pure LC Total Nucleic Acid Isolation Kit (Roche Life Sciences) by adding the 1 ml sample material (400µl DNase I treated sample material then diluted with 600µl PBS) directly into 1 ml of MagNA Pure Lysis and Binding-buffer (MPLB-buffer) following the field extraction method of Rosenstierne et al (2018). Briefly, sample and MPLB-buffer were mixed for lysis, before the magnetic glass particles (MGPs) were added and the contents mixed by gently flipping the tube. Using a small magnet, the MGPs were captured within the tube and washed three times in order to release bound nucleic acids. The MPLB-buffer contains guanidine isothiocyanate (GITCH) and detergent; which efficiently inactivates virus and proteins including the added DNase and thereby avoids heat or EDTA treatment.

### 2.3 Double-stranded cDNA synthesis and random amplification

For reverse transcription of viral RNA to cDNA and random amplification, we used the Whole Transcriptome Amplification (WTA) system in the REPLI-g Cell WGA & WTA Kit (Qiagen, Hilden, Germany). The kit includes four steps including genomic DNA removal, reverse transcription, ligation and random amplification with the phi29 DNA polymerase. We used two modifications of the protocol: firstly, the NA from the 10 µl input material remained bound to the MGPs from the NA extraction step (Rosenstierne et al., 2018). Secondly, instead of using the oligo dT primers provided in the kit for the reverse-transcription step, we used 20 µM 5′-phosphorylated random hexamers as described earlier (Rosenstierne et al., 2014).

### 2.4. Library preparation and Nanopore sequencing

Between 4 and 12 individual samples were multiplexed using the Rapid Barcoding Kit (Oxford Nanopore Technology). The DNA concentrations of the samples were determined using a Qubit Fluorimeter (Life Technologies) and used to normalize the input material for sequencing libraries. Here, sample concentrations were standardized to contain 400 ng DNA (in 7.5 µl) when possible, incubated with 2.5 µl of the individual barcodes for 1 minute at 30°C and then 80°C, before 10 µl of multiplexed samples were incubated for another 5 minutes at room temperature with the rapid adaptor enzyme. Samples were included despite not meeting the library input recommendations (400 ng). The protocol suggestion of a bead purification step when multiplexing four or more samples was omitted for faster library preparation with field use in mind. The libraries were loaded and sequenced on R9.4.1 flowcells on the MK1C device (Oxford Nanopore Technology) with default settings, fast base calling enabled and a run length of 20 hours. DNase/RNase free water was included as a negative control.

### 2.5. Data analysis

#### 2.5.1. Trimming

Reads were trimmed from barcodes using MinKNOW (version 21.02.1) before being imported to the Geneious Prime version 2021.2.2 (https://www.geneious.com) for data analysis. Using the quality trimming plugin tool BBduk2 (version 1.0), the raw reads were quality-trimmed with a minimum quality score of 7 at each end of each read as recommended for Oxford Nanopore reads by Geneious Prime and a read length above 150 nt.

#### 2.5.2. Metagenomic analysis of reads

The trimmed reads were blasted using Virosaurus (version 98, 2020_4.2), a curated offline database made for clinical metagenomics analysis, which contains full-length genome and segment (for segmented viruses) sequences from all known viral pathogens of vertebrates (Gleizes et al., 2021). HIV-1 was excluded to avoid the ethical dilemma resulting from randomized finds, which could not be acted upon. Only hits with an e-value at or below 1e-5, with a minimum length of 100 bp, a pairwise identity of ≥90% and ≥10 hits were considered valid.

#### 2.5.3. Mapping of reads and consensus sequence generation

The reads from hits identified above were mapped to the relevant reference sequence from Virosaurus (NC045512) using MiniMap2 for long-read assembly (Li, 2018) in Geneious Prime (version 2021.2.2.) for mapping and consensus sequence generation using default parameters. For typing SARS-CoV-2, a consensus sequence obtained from 15 of the then most commonly circulating variants in Denmark (B.1.617.2) was used as the reference genome sequence for the reads obtained after a full sequence run of 20 hours. Consensus sequences were generated using the most common base choice of SNP for the consensus sequence.

#### 2.5.4. Data analysis and visualization

Statistics and figures were performed using Microsoft Excel 2016 and GraphPad Prism version 8.3.0 for Windows (GraphPad Software, San Diego, California USA, www.graphpad.com).

## 3. Results

### 3.1. Evaluation of pretreatments on pooled samples

#### 3.1.1. Effect of pretreatments

We initially compared three pretreatment options: i) DNase treatment, ii) filtration, iii) DNase treatment followed by filtration, and no pre-treatment (as control) on the 16 pooled samples described in 2.1.1. As shown in Table 1, adding a pretreatment step to the metagenomic protocol increased the percentage of BLAST hits corresponding to SARS-CoV-2 from 31.8% (7/27) without any treatment to 99.9 % using DNAse followed by filtration. Furthermore, pretreatment lead to a much faster accumulation of SARS-CoV-2 mapped reads (Figure 1A and B). Without any pretreatment, too few SARS-CoV-2 BLAST hits to meet the quality requirements of >10 hits were obtained. With DNAse or filtration as individual pretreatments, SARS-CoV-2 hits did meet the quality requirements (Table 1) alongside BLAST hits for Orthohepevirus A (accession no.: MF444119).

**Table 1.**
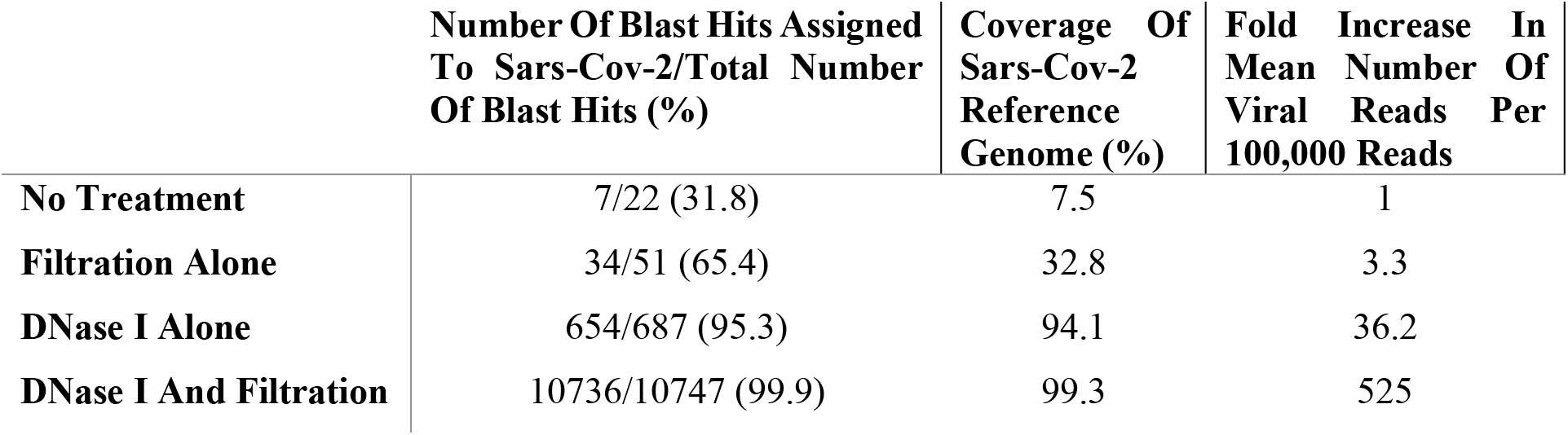
Pretreatment comparison to no treatment after 20 hours sequencing. No treatment BLAST hits value is also given.

**Figure 1.**
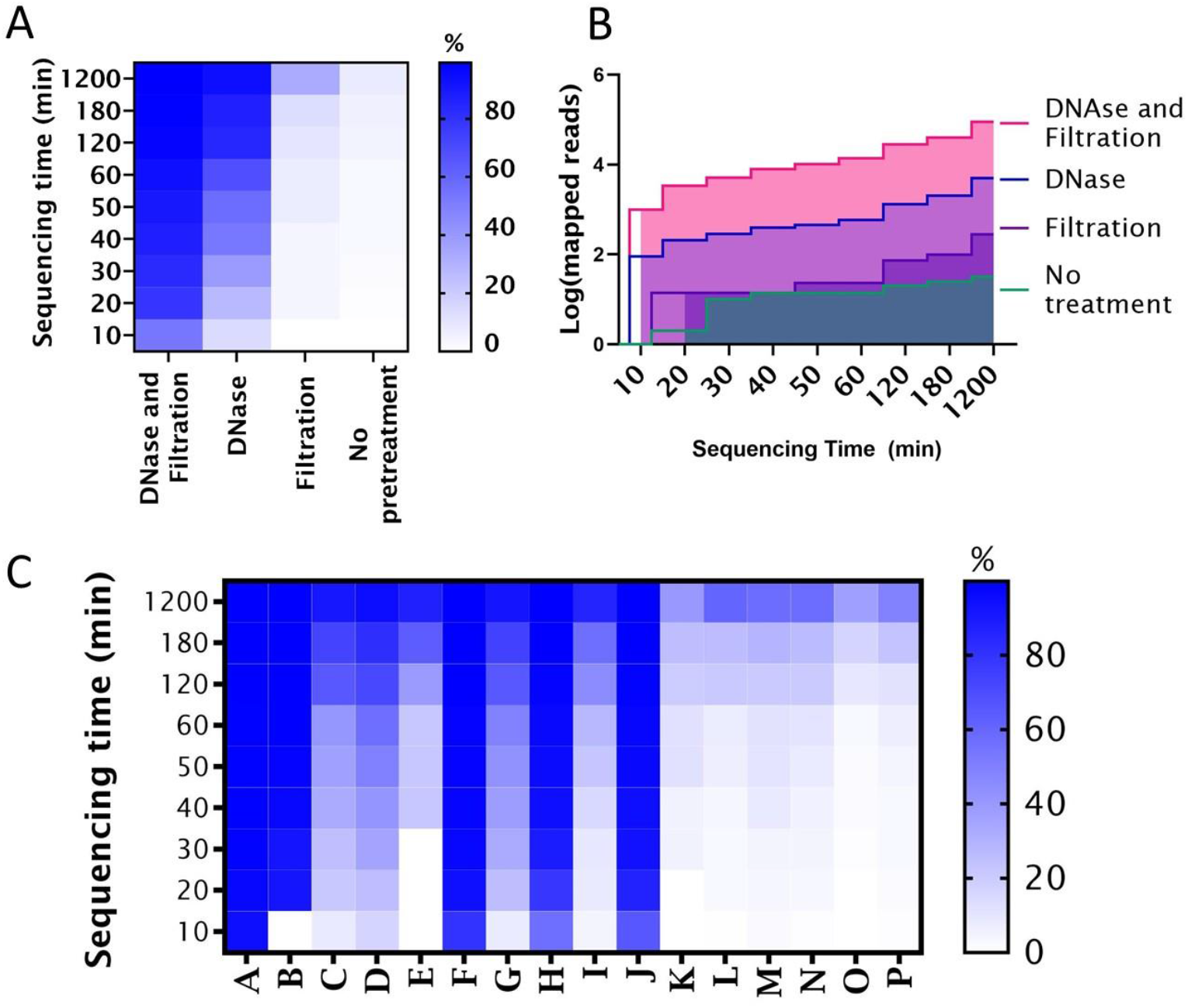
Influence of virus genome content and sample pretreatment on the number of SARS-CoV-2 sequence reads obtained. **Pooled samples:** A) Coverage of the SARS-CoV-2 NC045512 reference genome (%) for each evaluated pretreatment, B) Comparison of accumulated SARS-CoV-2 sequence reads over time for the indicated pretreatments. **Individual samples**: C) X axis: clinical samples (A-N) CT-values in the range of 18-36 and O being a negative swab and P being a water control (See also Table 1). Left Y axis: Sequencing time; Right Y axis: Percentage coverage of the SARS-CoV-2 (Acc. No. NC045512) reference genome (%).

#### 3.1.2. Assembling reads to reference genomes to spot false-positive BLAST hits

The reads of each pretreatment were assembled to pathogens that met the BLAST hits requirements as described in section 2.5.2. and visually inspected. Orthohepevirus A met the requirements for samples from the DNase alone and filtration alone as pretreatments. However, upon inspection all reads tiled at just two regions of 220 bp and 124 bp in the reference genome, respectively (genome coverage = 1.7% using either pretreatment) and were, therefore, disregarded as false-positive matches. This contrasted with the reads that mapped to the SARS-CoV-2 reference genome, which were evenly spread across the entire genome and were therefore regarded as true positive matches. The combination of DNase I and filtration pretreatments only needed 10 minutes of sequencing time before over 50% (53.3%) of the SARS-CoV-2 genome was covered. For comparison, using DNase I pretreatment alone, it took 40 minutes to obtain similar results with 52.5% of the reference genome covered.

### 3.2 Evaluation of pretreatment on individual samples

Using the same pretreatment conditions of DNase I treatment followed by filtration, 15 individual samples and one water control were analyzed. All samples with a CT-value at or below 30 achieved a high reference coverage (85.8-100.0%) after 1200 minutes accumulation compared to 40.0-60.5% for samples with a CT-value at or above 33 (Table 2).

**Table 2.** Read processing characteristics and genotype matching to Delta consensus sequence after 20 hours sequencing time of individual samples.

| Sample Name | SARS-Cov-2 CT-Value | Mapped Reads | SARS-Cov-2 Reads Pr. 100,000 Reads Generated | Fraction Of Reference Covered (%) | Clade | Lineage |
| --- | --- | --- | --- | --- | --- | --- |
| A | 18 | 1,188,028 | 92,459 | 100 | 21J (Delta) | AY.4 |
| B | 18 | 401,143 | 97,894 | 100 | 21J (Delta) | AY.4 |
| C | 21 | 14,267 | 5,277 | 91.3 | 21J (Delta) | AY.4.5 |
| D | 21 | 18,084 | 13,096 | 95.1 | 21J (Delta) | AY.43 |
| E | 24 | 3,648 | 63,921 | 87.3 | 21J (Delta) | AY.42 |
| F | 24 | 445,092 | 84,910 | 100 | 21J (Delta) | AY.29 |
| G | 27 | 8,543 | 2,043 | 92.1 | 21J (Delta) | AY.42 |
| H | 27 | 171,421 | 54,764 | 100 | 21J (Delta) | AY.33 |
| I | 30 | 3,248 | 1,783 | 85.8 | 21J (Delta) | AY.42 |
| J | 30 | 367,216 | 68,210 | 100 | 21J (Delta) | B.1.617.2 |
| K | 33 | 819 | 2,588 | 53.2 | 21J (Delta) | FAIL |
| L | 33 | 550 | 233 | 56.1 | 21J (Delta) | FAIL |
| M | 36 | 1,902 | 1,151 | 60.9 | 21J (Delta) | FAIL |
| N | 36 | 1,130 | 1,073 | 57.6 | 21J (Delta) | FAIL |
| O | >38 | 439 | 457 | 36.8 | 21J (Delta) | FAIL |
| P | H<sub>2</sub>O. No Ct value | 575 | 273 | 49.1 | 21J (Delta) | FAIL |

### 3.3 SARS-CoV-2 typing

Consensus sequences were made as described in section 2.5.3. and typed by Nextclade (version 1.10.0 available at https://clades.nextstrain.org/results) and Pangolin (version 3.1.16, lineages version 2021-11-25 available at https://pangolin.cog-uk.io/). To obtain most possible sequence data, reads were obtained after 1200 minutes. This showed that all 14 were identified as clade 21J (Delta), and typing by Pangolin identified various Delta sub-lineages for all samples with a CT-value below 33 (Table 2).

## 4. Discussion

On-site surveillance of emerging or re-emerging virus diseases is important to alert relevant public- and veterinary-health authorities to enable a fast response before substantial outbreaks develop. If such surveillance is to be implemented in resource-limited settings, then detection at point of incidence is needed. mNGS allows non-biased investigation and detection of viral agents which could circumvent the need for elaborate and highly specialized knowledge of symptoms and pathogens for specific assays which is often an obstacle because a wide variety of viral agents can cause similar symptoms and novel viruses cannot be anticipated. A single test system that detects diverse viruses in an unbiased manner will be important for future fast responses.

### 4.1 Comparison to other metagenomic protocols for virus detection; prospects for field-deployability of this protocol

One of the major challenges with unknown pathogen detection using mNGS is the low proportion of viral NA in a given clinical sample compared to the abundant non-viral components. This can consequently lead to the pathogen not being detected in an infected individual. A well-used strategy to overcome this problem is host-depletion with nuclease while filtration treatments are some of the most commonly used strategies alongside centrifugation (Hall et al., 2014). Others have reported that nuclease treatment alone generated a higher proportion of viral reads than a passage through a 0.45 µm filter followed by nuclease treatment (Rosseel et al., 2015). That study looked at virus in spiked serum and tissue samples using a protocol developed for metagenomic virus detection in a laboratory. The study used TURBO DNase, which has been used in several viral metagenomic studies (Greninger et al., 2015; Hall et al., 2014; Kafetzopoulou et al., 2019). However, the use of DNase Set I, as described here, halves the treatment time and avoids the incubation at 37°C in contrast to room temperature. Furthermore, the use of Illumina sequencing, which can generate more reads of a higher quality, is not feasible in the field. Greninger et al. (2015) demonstrated a metagenomic approach using the more portable Nanopore technology for RNA virus detection and this method has been used by others during disease outbreaks, e.g. for Lassa fever (Kafetzopoulou et al., 2019). However, this method requires a thermal cycler. By switching to isothermal reactions in a simpler incubator, like for this protocol, a mNGS protocol could become less complex and more suitable for less specialized mobile laboratory systems and even field-testing. Furthermore, the library preparation for Nanopore sequencing used by Greninger et al. (2015) includes many “hands-on” steps (e.g., three magnetic purification steps) whereas the Rapid Barcoding kit used here has the advantage of not requiring purification steps plus quick fragmentation and adapter ligation. Using the similar Rapid Sequencing kit, the concentration normalization step can also be avoided when sequencing only single samples. The protocol presented in this study has been developed with the aim of excluding large non-mobile advanced equipment such as centrifuges, extraction robots, thermal cyclers and sequencing robots (e.g., the Illumina apparatus) in an attempt to introduce simpler and mobile metagenomic pathogen detection.

### 4.2 Sensitivity and evaluation of individual samples’ results

Like any diagnostic assay, the efficiency of mNGS as a diagnostic tool depends strongly on the amount of virus nucleic acid in the sample (Rosseel et al., 2015). Thus, the higher viral loads at the start of the symptomatic infection would facilitate detection by mNGS because the amount of viral NA will be larger. For the protocol described here, the CT-values of the samples post field NA extraction were higher than for the same samples when immediately tested at the TCDK the same day (data not shown). However, by removing free-floating DNA with DNase and filtering away larger components like host-cells, bacteria and other interfering contaminants, the proportion of SARS-CoV-2 RNA derived reads increased enough to be detected readily in samples. The drop in sensitivity might be explained by this protocol selecting only for extracellular viral RNA whereas the one-step RT-qPCR, as used by the TCDK, allows for both intra- and extracellular viral RNA quantification. The inclusion of the Repli-G WTA step for random amplification is thus an attempt to outweigh this loss of sensitivity. Another pitfall when using mNGS for pathogen detection is the risk of false-negative and false-positive results and it is therefore important to have non-template controls included such as water (Simner et al., 2018). There are no accepted golden guidelines for proper mNGS protocols for virus detection yet. Therefore, defining a cut-off for a negative result (absence of pathogen) needs to be empirically determined. Here, reads were observed in negative swabs and water controls in accumulated reads after 1200 minutes. Since this could have been caused at one or several steps during the procedure, e.g., as cross-contamination during the sample processing, from the environment or from extraneous sources of DNA from reagents used in the workflow. We therefore used a SARS-CoV-2-specific one-step qPCR assay to monitor contaminants after the NA extraction and after the random amplification using the phi29 DNA polymerase. The reverse transcriptase was omitted for the post random amplification step to look only for ds-cDNA from SARS-CoV-2, which would have been generated following the Repli-G WTA protocol and subsequently be sequenced. In duplicate qPCR assays, no positive signals were detected for the water control nor for negative swab following the one-step qPCR assay but showed a clear amplified signal (lower CT-values) for SARS-CoV-2 in clinical positive samples after random amplification (data not shown). A possible explanation seems therefore to be that this occurred during demultiplexing. In this study, we find that our method reliably allows for the retrieval of Pangolin typing-grade sequences when the CT-value was below 30, whereas this could not be obtained with higher CT-values, although SARS-CoV-2 clades were still identified (Table 2).

### 4.3 Future investigations

A limitation of our study is that our test of concept was only performed using a particular RNA virus, namely SARS-CoV-2. Testing for other RNA viruses and DNA viruses, albeit without the reverse transcription step, is ongoing to establish a more generally applicable virus detection method. Furthermore, it is of relevance to investigate whether the protocol, when used on serum and tissue samples, produces less viral reads than pretreatment with DNase I, as described by Rosseel et al (2015). Our field extraction method using MPLB-buffer was originally developed for whole blood samples and urine (Rosenstierne et al., 2018). With this study, the field method has also proven useful for throat swab materials. For mNGS on tissue samples, the hand extraction protocol might need adjustments, which remains to be investigated and possibly optimized. Another future possibility is to further improve the bioinformatical pipeline, so the workflow becomes faster. The use of pretreatments like DNase I or filtration alone did provide some spurious BLAST hits that fitted within the initial quality requirements especially for Orthohepevirus A but were removed upon closer inspection of mapped reads to the Orthohepevirus reference (accession no.: MF444119). This shows that database searches can be misleading for non-trained personal. However, this was not relevant for this study since 99.9% of the BLAST hits were on SARS-CoV-2, but this might change with different sample materials or for other viruses. One approach could be to circumvent the BLAST step and simply assemble the quality trimmed reads directly onto all known virus genomes; however, this database needs to be built with careful selection and continuous updating.

## 5. Conclusions

The world is regularly experiencing emerging zoonotic RNA viruses in animals and humans, most recently the pandemic RNA virus SARS-CoV-2. For rapid detection in a field setting, the response-time to the relevant stakeholders needs to be decreased and equipment should be transportable. By introducing a 15-minute pretreatment, with DNase I treatment followed by a 0.22µM filtration step, the field-deployable protocol with Nanopore sequencing technology greatly increased the probability of detecting SARS-CoV-2 in a clinical sample with a CT-value below 33. The dramatic increase in the proportion of generated viral reads obtained in the subsequent Nanopore sequencing allowed for a simple BLAST identification of SARS-CoV-2 with even coverage across the genome when mapped to a reference sequence, which enabled identification of the virus clade and often the lineage. This principle may be useful in simple laboratory settings or for field identifications at the point of incidence for virus pathogens in animal and human swab samples.

## Data Availability

Data produced in the present study are available for research upon reasonable request and permission from The Danish Health Data Authority

## CRediT authorship contribution statements

**Anna S. Fomsgaard:** Conceptualization, methodology, investigation, formal analysis, writing.

**Morten Rasmussen:** Conceptualization, review of results and manuscript.

**Katja Spiess:** Conceptualization, review of results and manuscript.

**Anders Fomsgaard:** Conceptualization, methodology, review of results and manuscript.

**Graham J. Belsham:** Supervision, review of results and manuscript.

**Jannik Fonager:** Supervision, conceptualization, methodology, investigation, formal analysis, writing.

## Acknowledgements

Thank you to Sofie Holdflod Nielsen and Arieh Cohen from “Test Center Danmark” for supply of relevant samples.

This work was done as part of the TELE-Vir project, supported by funding from the European Union’s Horizon 2020 Research and Innovation program under grant agreement No 773830: One Health European Joint Programme.

